# Mixed Methods Sequential Epidemiological Protocol (MMSEP) for Evaluation of Time Utilization by the Community Health Workers and Factors Affecting its Optimization

**DOI:** 10.1101/2022.07.20.22277875

**Authors:** Poonam Sanjay Kalne, Shiv Hiren Joshi, Pooja Sanjay Kalne, Ashok Madhukar Mehendale

## Abstract

**Background:** Many countries have a significant shortage of healthcare staff, resulting in a reduction in the range and quality of services. Low and middle-income countries rely heavily on Community Health Workers. Existing data suggests a lack of clarity regarding the scope of work as well as about the utilization of the working hours. The purpose of this work is to propose a protocol to evaluate the time utilization of the Community Health Workers and to identify the promoters and challenges that they come across while delivering their duties so that the workflow optimization processes can be hypothesized.

**Methods:** The Mixed Methods Sequential Epidemiological Protocol incorporates three phases. Phase I includes free listing to identify Community Health Workers’ workflow using a generic form with a single open-ended question for each cadre. After pile-sorting the free lists, the pile names should be used as categories for the time-motion study to be done in phase II. For each of these categories, time spent should be recorded and analyzed. Phase III includes in-depth interviews with Community Health Workers who use considerably varied times for the same category of work.

**Discussion:** The intended study will help in comparing the workflow of different cadres of health workers and finding out different factors promoting and inhibiting better utilization of their working hours. This approach will inform reasonable and feasible improvements to health-related planning of services and utilization of the health workforce and will help in increasing the efficiency of health workers to deliver services.

**Author contributions:** 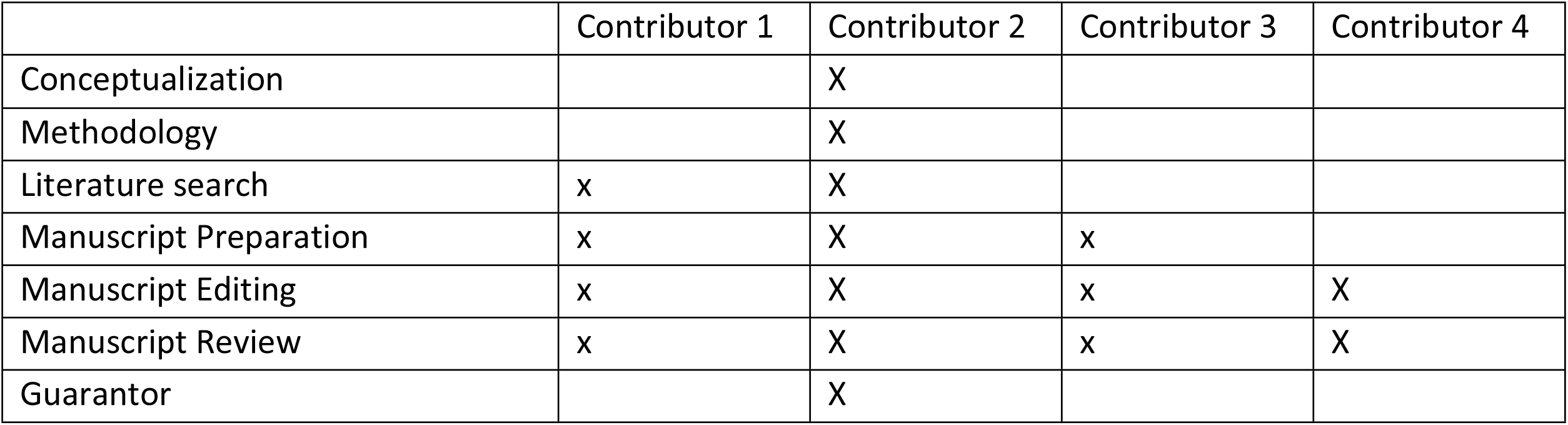

## Introduction

The CHWs (Community Health Workers) play a vital role in the health care system due to their outstanding community-based services that cater the rural community. High neonatal and maternal mortality in India is thought to be due to inadequate infrastructure, poor health services, and lack of human resources which may be attributed to a meagre health budget that accounts for less than 2% of government spending as a percentage of GDP (1). Such economic constraints necessitate the need to study the workflow of CHWs to identify the related promoters and to target the challenges faced by them to optimize their workflow and improve the overall status of healthcare delivery in the country.

CHWs bridge the gap between the people and the healthcare system of the country to improve the health related outcomes in the domains of maternal health, child health, nutrition, communicable diseases, non-communicable diseases and immunization (2). Healthcare systems, particularly from the low and middle-income countries, rely heavily on CHWs (3). These are individuals from the community who are chosen by the community to assist the health system in providing primary health care within their community. Moreover, most of the CHWs are volunteers (3). According to the Shrivastav Committee’ s recommendations (1975), the Rural Health Scheme was created by the Indian government in 1977 with the purpose of “putting people’ s health in people’ s hands (4).” At the International Health Conference of Alma Ata in 1978, primary health care was chosen as a method for achieving the WHO’ s global, societal goal of “Health for All by 2000AD (5,6).” Many low- and middle-income countries do not have enough resources to sustain and train the literate health workforce that is needed to provide quality healthcare services to the community. As a result the burden of delivering these services is shared by the different cadres of CHWs (7). Accredited social health activist (ASHA), Anganwadi workers (AWW), and Auxiliary Nurse Midwives (ANM) are examples of the various cadres under CHWs. Existing evidence suggests that the high coverage of CHW visits increases the household awareness and brings about the change in behaviour of the people towards their health-related practices.

The CHWs have been increasingly recognized as the key to attaining overall improvement in the health status of a country (6). According to the existing data, there is a lack of clarity on the actual scope of work-related activities of ANMs, ASHAs, and AWWs. It is necessary to understand their existing workflow to develop tangible work optimization processes. To identify the social, organizational, and interpersonal factors that either interrupt or facilitate the CHWs programs (5), a comprehensive protocol for evaluation of their workflow is needed. Hence, a mixed methods sequential epidemiological protocol (MMSEP) is proposed to evaluate the time utilization and the related promotors and challenges to provide necessary health services to the deserving population of society within time (8,9).

The MMSEP is guided by the following research questions:

Research Question 1 -What is the time use pattern of CHWs?

Research Question 2 - Which factors facilitate better utilization of working hours of CHWs?

Research Question 3 - Which factors prevent better utilization of working hours of CHWs?

## Methods

The Mixed Methods Sequential Epidemiological Protocol:

This evaluation will be conducted in 3 phases (Figure 1):

**Figure 1.**
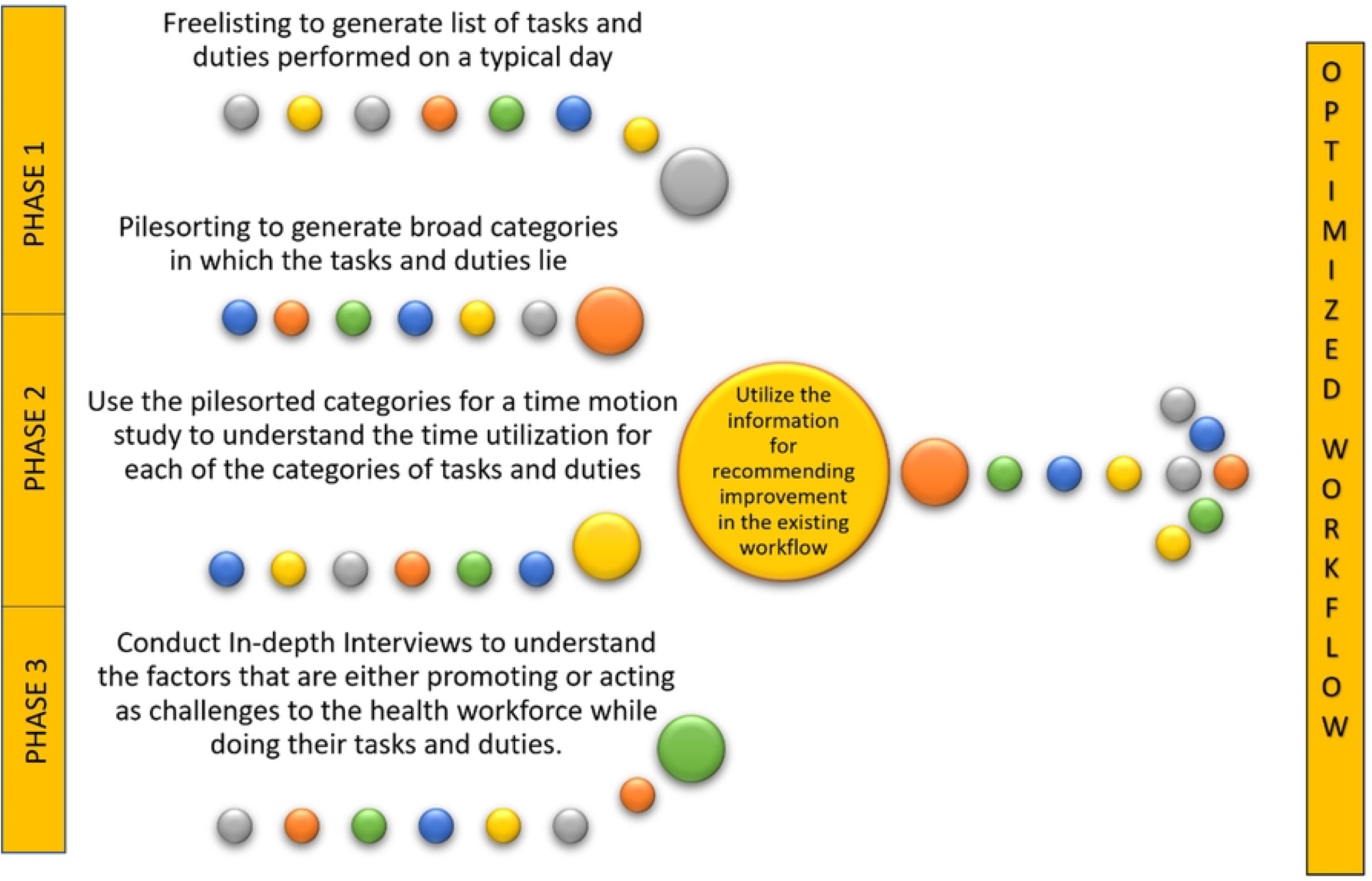
Summary of the flow of events under the MMSEP

### Phase I

In the first phase, freelisting will be done to identify the activities which are a part of the workflow of a typical day. The freelist technique is used to elicit the elements or members of a domain. For domains that have a name or are easily described, the technique is very simple: the researcher just needs to ask a set of respondents to list all the members of the domain. Freelisting is a qualitative interviewing technique that works well for involving communities and uncovering shared objectives. It is a great tool for quickly examining how people define and think about a certain health-related subject. The domain of interest in the MMSEP is the “Duty related activities”. Freelisting should be done separately for each cadre of the CHWs. These replies should be collected using a generic freelisting form with a single open-ended question. We recommend the question, “What activities are part of your workflow on a typical day?” Each respondent should be urged to think of as many activities as they could, and they should be encouraged to include even the most insignificant ones. After the freelisting is completed, the pilesorting should be done. Pilesorting is an ethnographic technique used to provide insight into how people conceptually organize information and relationships between items (10). The free pilesort technique is used primarily to elicit from respondents’ judgments of similarity among items in a domain. This technique begins with a stack of blank cards. The unique “duty related activities” that were identified from the freelists are mentioned on each of these cards hence giving us a stack of the same number of cards as the identified unique “duty related activities”. This stack of cards is shuffled randomly and given to the researcher with the following instructions: “Here are a set of cards representing ‘duty related activities’ of ASHAs (This can be replaced with the desired name of the cadre that is under assessment). I would like you to sort them into piles according to how similar they are. You can make as many or as few piles as you like.” The pilesort exercise should be repeated with at least 30 independent individuals identified by the researcher.

### Phase II

After analysing the results of phase I and pile-sorting the free lists, the pile names shall function as categories which can be utilized for the time-motion study. The time-motion study is a process in which the duration of each activity performed by a subject is noted to build the workflow, ensure efficiency and effectiveness, and reduce wastage of resources. For each of the categories, the time spent by the different cadres of CHWs should be noted and then analysed.

### Phase III

In-depth interviews should be organized with the CHWs who give significantly different times for the same category of the work to understand different factors that promote or create challenges in their workflow. An in-depth interview guide may be used for this purpose. With the help of in-depth interview, interviewers have more opportunities to probe for more information, ask follow-up questions, and return to important issues later in the interview to produce a rich understanding of attitudes, perspectives, and motivations.

### Data analysis plan

#### For Phase I

The data obtained from the free lists should be used to create an APAC formatted text file. This text file is to be imported into ANTHROPAC for calculating Smith’ s Cognitive salience. This shall be done separately for the free lists obtained from each of the cadres of the CHWs. The unique items identified from the free lists should be used for pile sorting. The pilesorting data can be further analysed by tabulating the item-by-item proximity matrix where each cell represents the proportion of the individuals who grouped each item pair together. A non-metric multidimensional scaling (MDS) can be used to identify the items that are closer to each other on the MDS plot. Such items are more likely to be grouped together as compared to the items that are at a greater distance from each other. Hierarchical cluster analysis can be used to objectively identify the clusters which are more likely to be made into distinct pile titles.

#### For Phase II

After pile-sorting, the identified categories (pile titles) should be noted. The data obtained from the time-motion study should be summarized using descriptive statistics to include the meantime with standard deviation for each category. Comparison of the time spent in each category and the time spent by different cadres for the same category of work should be noted. It is important to check for the distribution of the gathered data to compare means or medians, and accordingly a t-test or a Mann Whitney test can be applied. The mean working hours should also be reported. In an instance where some CHWs did not conduct the tasks freelisted by the other CHWs of the same cadre, their observation of time utilized should be excluded from the analysis while reporting to prevent the skewing of the findings.

#### For phase III

Following the in-depth interview, a transcript can be made in English. Thematic contents analysis can be done using the RQDA package of R version 4.1.1 to analyze these transcripts. The approach described by Braun and Clarke should be utilized to perform a thematic contents analysis of the transcripts (11). This method is selected to explore the views of the study participants using a well-suited and analytical approach. The researchers should familiarize themselves with the content of the transcripts by a thorough reading and codes should be created for the content. Each of the researchers should independently develop these codes and grouped them to generate themes. The themes should be identified and revised till a consensus is reached. After the first round of in-depth interviews, an interim thematic content analysis should be performed to set a baseline of findings. Following this, another set of interviews should be conducted and analyzed. This process should be continued till the researchers reach a point of data saturation meaning that no new findings are observed in the subsequent set of interviews upon carrying out an interim analysis.

## Discussion

Human resource in any sector of development is invaluable and more in the domain of healthcare delivery. Having an in-depth understanding of how the CHWs spend their time in the different domains of work-related activities can help to identify those domains which demand more time compared to the other domains. It also helps to identify the factors affecting these time utilization patterns both within the cadre and between the cadres. Such insight has direct implications on improvement of the work performance and to develop mechanisms for optimal utilization of the time of our limited healthcare workforce. Thus, this three-phased methodology of the protocol will be useful to guide the development, implementation and evaluation of future scaling-up strategies to accelerate change towards more sustainable health and care systems. Combining the time-motion study data with findings from free listing and in-depth interviews facilitate to develop an insight into healthcare workflow, inefficiencies, patient safety, and quality which may have significant implications (9,12).

### Strengths and limitations of the protocol

The MMSEP enables to quantify time utilization to provide medical services to the community. Recognizing how the CHWs use their time is a critical impediment to maximizing the benefits of community services. A mixed-method sequential approach will aid in the in-depth understanding of CHW’ s outputs, incentives, motivations, facilitators, and obstacles while providing healthcare services. The MMSEP can potentially aid in the improvement of processes and the optimization of CHW workflow and in turn their overall performance, as well as it will facilitate the development of new approaches to increase their work efficiency. It may include the risk of observers interfering with the work and time utilization patterns by their presence.

### Ethics and Dissemination

The use of this mixed methods epidemiological approach was approved by the Institutional Ethics Committee of Datta Meghe Institute of Medical Sciences (Deemed to be University) (Regd. No. ECR/440/Inst/MH/2013/RR-2019) on 31/01/2022 through their letter number DMIMS(DU)/IEC/2022/691. While implementing this methodology, the researchers shall involve the study participants after taking their written informed consent. Only after all the queries of the study participants are resolved, the researchers should start the data collection.

Further, the study participants should be made aware that they can choose to remove themselves from the study or a part of the study at any point in time and that it is not binding for them to continue participating in the study once they have given their consent. While analyzing and storing the collected data, the researchers should anonymize and delink the data. Only the principal investigator shall have access to the code that links the participant information to their identifiers. While all the interviews should essentially be audio-recorded, it should be done with the consent of the participants. Also, the participants shall be made aware that they can choose to go off the audio record if they wish for a part of the discussion and that whatever duration of audio is recorded, it shall stay confidential and it shall be accessible via a password protected drive only to the principal investigator. After the audios from the in-depth interviews are transcribed, researchers should take it back to the participants for ensuring the consistency of meaning and to check if the language and tone of describing the context is acceptable to them. These in-depth interviews shall be conducted at a place of convenience for the CHWs (ASHA, AWW, ANM) where their privacy can be ensured.

## Conclusion

The performance of community health professionals is becoming increasingly popular (CHWs) around the world; nevertheless, there are gaps in the research on CHWs’ involvement in community participation and empowerment. This study proposes a mixed methods sequential epidemiological protocol that may potentially be useful in creating an in-depth understanding of the time utilization patterns of the CHWs by using a phase wise evaluation of the workflow using established techniques in epidemiology to generate a hypothesis regarding the reasons causing sub-optimal utilization of time and how it may be improved.

## Data Availability

No datasets were generated or analysed during the current study. All relevant data from this study will be made available upon study completion.

## Acknowledgements

The researchers acknowledge the role of Community health workers who are instrumental to the healthcare delivery which is even more important for a developing country given the context of limited resource settings.

